# Upper and Lower Respiratory Tract Compartmentalization in Pediatric Stem Cell Transplantation

**DOI:** 10.1101/2025.10.15.25336763

**Authors:** Erica M. Evans, Madeline Y. Mayday, Emma M. Pearce, K Iwanaga, NP Ly, GD Church, Gustavo Reyes, Miriam R. Simon, Jacob Blum, Hanna Kim, Jessica Mu, Jazmin Baez-Maidana, Jeffrey J. Auletta, Peter J. Shaw, Erin M. Kreml, Paul L. Martin, Christine N. Duncan, Courtney M. Rowan, Kamar Godder, Caitlin Hurley, Geoffrey D.E. Cuvelier, Muna Qayed, Hisham Abdel-Azim, Amy K. Keating, Julie C. Fitzgerald, Rabi Hanna, James S. Killinger, Janet R. Hume, Troy C. Quigg, Paul Castillo, Prakash Satwani, Theodore B. Moore, Christopher C. Dvorak, Matt S. Zinter, the Pediatric Transplantation and Cell Therapy Consortium

**Affiliations:** Division of Critical Care Medicine, Department of Pediatrics, University of California, San Francisco, San Francisco, CA, USA; Departments of Laboratory Medicine and Pathology, Yale School of Medicine, New Haven, CT, USA; Division of Pulmonary Medicine, Department of Pediatrics, University of California, San Francisco, San Francisco, CA, USA; Hematology/Oncology/BMT and Infectious Diseases, Nationwide Children’s Hospital, Columbus, OH, USA; CIBMTR® (Center for International Blood and Marrow Transplant Research), National Marrow Donor Program/Be The Match, Minneapolis, MN, USA; The Children’s Hospital at Westmead, Westmead, NSW, Australia; Department of Child Health, Division of Critical Care Medicine, University of Arizona, Phoenix, AZ, USA; Division of Pediatric and Cellular Therapy, Duke University Medical Center, Durham, NC, USA; Harvard Medical School, Boston, Massachusetts; Division of Pediatric Oncology, Department of Pediatrics, Dana-Farber Cancer Institute and Boston Children’s Hospital, Boston, MA, USA; Indiana University, Department of Pediatrics, Division of Critical Care Medicine, Indianapolis, IN, USA; Cancer and Blood Disorders Center, Nicklaus Children’s Hospital, Miami, FL, USA; Division of Critical Care, Department of Pediatric Medicine, St Jude Children’s Research Hospital, Memphis, TN, USA; CancerCare Manitoba, Manitoba Blood and Marrow Transplant Program, University of Manitoba, Winnipeg, Manitoba, Canada; Alberta Childrens Hospital, Calgary, Alberta, Canada; Aflac Cancer & Blood Disorders Center, Children’s Healthcare of Atlanta and Emory University, Atlanta, GA, USA; Department of Pediatrics, Division of Hematology/Oncology and Transplant and Cell Therapy, Keck School of Medicine, University of Southern California, Los Angeles, CA, USA; Loma Linda University School of Medicine, Cancer Center, Children Hospital and Medical Center, Loma Linda, CA, USA; Center for Cancer and Blood Disorders, Children’s Hospital Colorado and University of Colorado, Aurora, CO, USA; Department of Anesthesiology and Critical Care, Perelman School of Medicine, Children’s Hospital of Philadelphia, University of Pennsylvania, Philadelphia, PA, USA; Department of Pediatric Hematology, Oncology and Blood and Marrow Transplantation, Pediatric Institute, Cleveland Clinic, Cleveland, OH, USA; Division of Pediatric Critical Care, Department of Pediatrics, Weill Cornell Medicine, New York, NY, USA; University of Minnesota, Department of Pediatrics, Division of Critical Care Medicine, Minneapolis, MN, USA; Pediatric Blood and Marrow Transplantation Program, Texas Transplant Institute, Methodist Children’s Hospital, San Antonio, TX, USA; Section of Pediatric BMT and Cellular Therapy, Helen DeVos Children’s Hospital, Grand Rapids, MI, USA; University of Florida, Gainesville, UF Health Shands Children’s Hospital, Gainesville, FL, USA; Division of Pediatric Hematology, Oncology and Stem Cell Transplantation, Department of Pediatrics, Columbia University, New York, NY, USA; Department of Pediatric Hematology-Oncology, Mattel Children’s Hospital, University of California, Los Angeles, CA, USA; Division of Allergy, Immunology, and Bone Marrow Transplantation, Department of Pediatrics, University of California, San Francisco, San Francisco, CA, USA

**Keywords:** Hematopoietic Stem Cell Transplantation, Intensive Care Units, Pediatric, Immunocompromised Host, Bronchoalveolar Lavage, Transcriptome

## Abstract

**Rationale:** Lung injury after hematopoietic stem cell transplantation (HCT) occurs due to infection, chemotherapy toxicity, and alloreactive inflammation. Analyses of bronchoalveolar lavage (BAL) fluid have revealed dominant pathobiologic signatures, but minimally-invasive diagnostics are needed.

**Objectives:** To determine whether microbiome and gene expression perturbations are shared along the respiratory tract or isolated to the alveoli in pediatric HCT patients with lung injury.

**Methods:** We performed bulk RNA sequencing on 189 paired nasal and BAL samples from 160 patients enrolled at 28 children’s hospitals (2016-2021). Microbial and human transcripts were compared using multivariable models accounting for age, sex, and paired sampling.

**Measurements and Main Results:** BAL and nasal transcriptomes differed across 13,698 genes, 48 cellular components, and network interactions linking inflammation, reactive oxygen species, and immunometabolism. Minimal BAL-nasal correlation was observed in gene expression levels (median ρ=0.03, IQR –0.03 to 0.08) or fractional abundance of key cells such as neutrophils and CD8+ T-cells. BAL microbiomes harbored fewer commensal bacteria and more fungi and DNA viruses. BAL bacterial RNA was associated with diminished immune signaling whereas nasal bacterial RNA aligned with inflammatory gene expression. Further, only BAL microbial RNA was linked to transcriptional shifts in epithelial injury response, keratinization, and collagen deposition. Finally, BAL commensal microbiome depletion, epithelial injury, and immune dysregulation signatures were associated with death or ≥7 days of mechanical ventilation in 30% of patients, whereas nasal samples provided minimal prognostic information.

**Conclusions:** These data support alveolar compartmentalization in pediatric HCT and emphasize the ongoing need for minimally-invasive but informative diagnostics.

## INTRODUCTION

Children with leukemia, hemoglobinopathies, and immunodeficiencies refractory to medical treatment may be cured by hematopoietic stem cell transplantation (HCT), which employs high-dose chemoradiation followed by intravenous infusion of a donor cellular graft to reconstitute blood cell lineages (1). While outcomes have improved in recent decades, lung injury still occurs in 25-40% of patients due to infection, chemotherapy toxicity, and alloreactive inflammation, which combine to accelerate pulmonary dysfunction and tissue injury (2–4). Together these triggers lead to intensive care admission in 15-20% of patients and 50-60% mortality when mechanical ventilation is required (5, 6). Therefore, strategies to mitigate pulmonary complications of HCT are greatly needed (7, 8).

To investigate the pathobiology of post-HCT lung injury, the *Pediatric Transplant and Cell Therapy Consortium* enrolled 229 patients undergoing 278 clinically-indicated bronchoscopic procedures for evaluation of post-HCT pulmonary complications across 32 children’s hospitals in the United States, Canada, and Australia (9). Using metatranscriptomic sequencing of bronchoalveolar lavage (BAL) fluid, we identified 4 subtypes of lung injury defined by varying patterns of dysbiosis, infection, inflammation, and cellular injury. This illuminated previous clinical diagnoses such as Idiopathic Pneumonia Syndrome (IPS) by enhancing pathogen detection and defining the principal pathobiologic perturbations in the lung. In addition, by linking alveolar biology to clinical outcomes, we provided an empirical framework for risk stratification in this population. For example, patients with depleted pulmonary microbiomes, diminished alveolar macrophage activity, activated T-cell signaling, and signatures of epithelial mesenchymal transition had 3– to 4-fold higher in-hospital mortality than their counterparts.

However, BAL is an invasive procedure that is not well-suited to repeat sampling; a minimally-invasive sampling method of the upper respiratory tract that recapitulates lower airway biology could improve timely diagnosis and facilitate closer disease monitoring (10). In support of this premise, the upper and lower respiratory tracts are frequently exposed to the same stimuli, whether respiratory virus, aspiration, or chemotherapy exposure, and communicate closely via paracrine and hematogenous signaling mechanisms (11–14). However, the respiratory tract also features a proximal-to-distal gradient of highly specialized histology, epithelial cell function, immunology, and microbiology (15–17). Therefore, it remains unknown whether upper respiratory samples can suitably approximate lower respiratory biology in research and clinical settings.

To directly address this knowledge gap, we analyzed paired BAL and nasal swabs collected simultaneously from patients in the study described above. By contrasting transcriptomes and microbiomes across the upper and lower respiratory tracts, we evaluated shared and distinct signaling patterns and clarified the utility of nasal swabs as a minimally-invasive surrogate for BAL in the HCT population. Some of the results have been previously reported in the form of an abstract (18).

## METHODS

### Patients

As previously described, from 2014-2022, 32 children’s hospitals in the United States, Canada, and Australia screened HCT recipients for clinically-indicated BAL procedures. Eligible patients or their guardians consented to biobanking of leftover BAL fluid (BALF) and a subset consented to collection of a paired nasal swab. Clinical care, including microbiologic testing, antimicrobial treatment, and immunomodulation, was not standardized and performed at the best judgment of each hospital. Patient follow-up data were collected through hospital discharge.

### Samples

BAL was performed by pediatric pulmonologists using institutional protocols. After aliquoting BALF for clinical testing, excess lavage was aliquoted into sterile cryovials. Anterior nasal swabs were obtained during the BAL procedure using sterile Floq-swabs. All samples were placed immediately on dry ice and stored at –70°C until processing.

### Data Generation

As previously described (9, 19), 200uL of BALF underwent mechanical homogenization in RNA-preserving media followed by column-based RNA extraction (Zymo) and non-directional library preparation (NEB Ultra II). For this study, nasal swabs were combined with 1mL RNA-preserving media and vortexed to release mucous, followed by bead-based mechanical homogenization, RNA extraction, and sequencing library preparation as above. Both nasal and BALF RNA were sequenced on Illumina NovaSeq instruments and resultant.fastq files were processed on the CZID pipeline v7.1 to produce human and microbial transcript counts. Human transcript counts were normalized to sequencing depth and variance; microbial transcript counts were normalized to a spike-in control to approximate RNA mass and further adjusted for background contamination found in negative controls (20).

### Analysis

BAL and nasal transcriptomes were compared graphically using MDS plots and normalized counts were modeled against body site, peripheral blood immune cell counts, microbe masses, and clinical outcomes using generalized estimating equations accounting for age, sex, and repeat samples from the same patient (21). Differentially expressed genes underwent pathway enrichment using MSigDB, Reactome, and GOBP datasets (22). BAL and nasal transcriptomes were compared within patients using Spearman rank-based correlation of normalized gene counts. BAL and nasal cell types were estimated using CIBERSORTx with recent reference cell atlases (23–25). Analyses were repeated at the microbial level by substituting human gene expression for microbial masses. All analyses involving ≥10 comparisons underwent FDR-adjustment of p-values. Finally, BAL and nasal gene expression and taxonomic data were modeled together using machine learning followed by dimensionality reduction and clustering at the patient level (*MOFA*) (26). Detailed methods are provided in the **Online Supplement.**

## RESULTS

### Patients

We collected 189 paired nasal swabs and BAL samples from 160 patients at 28 children’s hospitals (**Figures 1A, 1B**). Patient demographics varied broadly and most underwent HCT for malignant conditions using a variety of allograft donors and preparative regimens (**Table 1**). Sample collection occurred a median 120 days post-HCT (IQR 39-371) at which point respiratory symptoms had been present for a median 8 days (IQR 3-22) and half the cohort required supplemental oxygen preceding BAL. Many patients had poor immune function at enrollment, as evidenced by nearly 50% rate of GVHD and a median blood ALC of 0.46 cells/mL (IQR 0.18-1.12). After sample collection, approximately half of cases were adjudicated as infection and half as IPS. Intensive care was required in over 50% of cases, and 30% died or required ≥7 days of mechanical ventilation.

**Figure 1.**
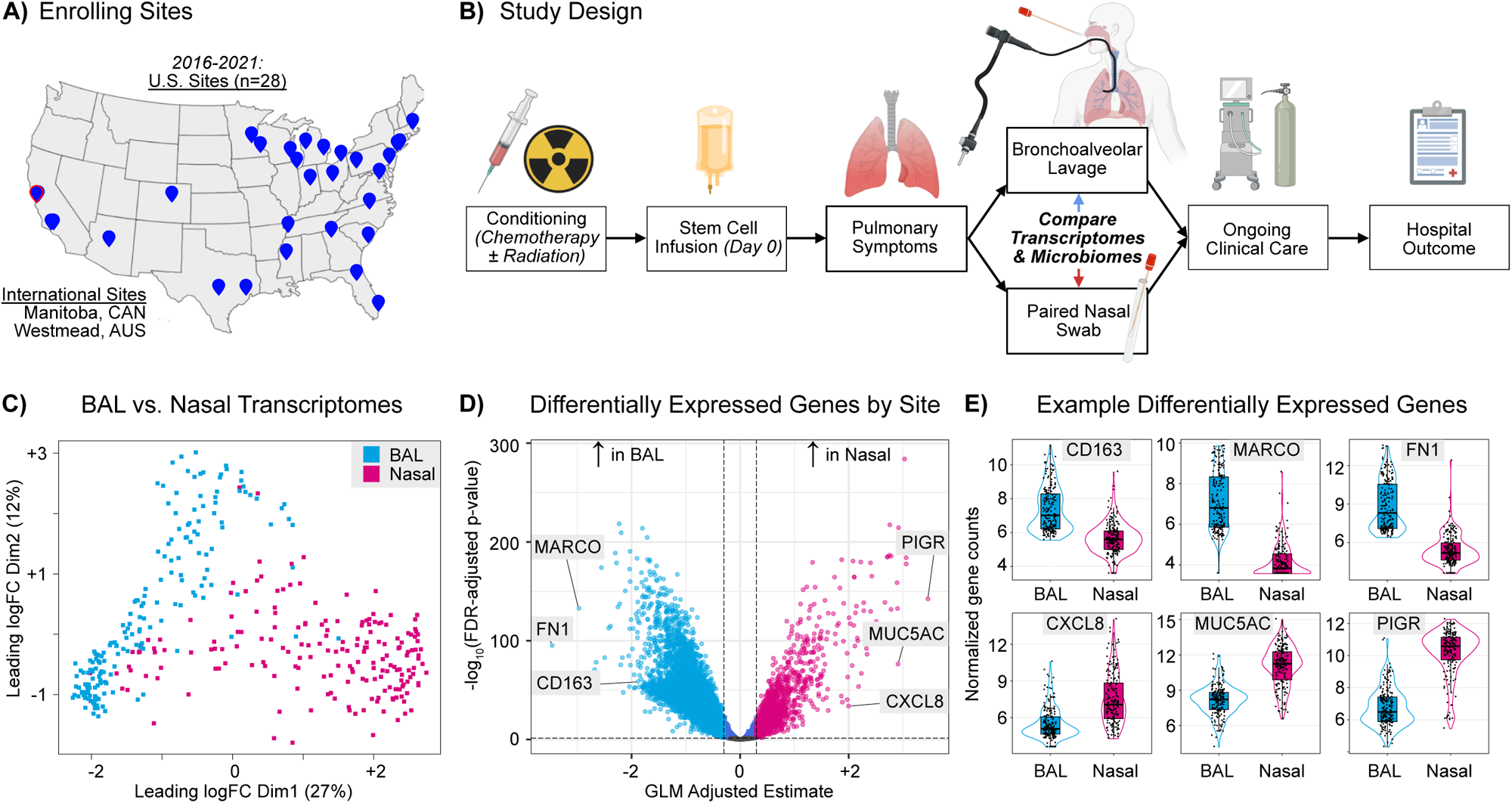
**Legend: (A)** Patients were enrolled from 28 children’s hospitals in the United States, Canada, and Australia between 2016-2021. **(B)** As part of clinical care, all patients received conditioning chemotherapy followed by autologous or allogeneic stem cell infusion on transplant day 0. All patients developed pulmonary symptoms such as cough, dyspnea, and/or hypoxia and underwent clinically-indicated bronchoalveolar lavage as part of diagnostic evaluation of post-HCT lung injury. Need for BAL was determined by the clinical team. Leftover BAL sample and a nasal swab were collected with informed consent and patients were followed through hospital discharge. **(C)** BAL and nasal gene counts were normalized to sequencing depth, sample distances were calculated and underwent multi-dimensional scaling (MDS), and samples were plotted according to the top two dimensions (x– and y-axes). Blue shading indicates BAL samples and pink shading indicates nasal samples. Samples closer in two-dimensional space are more similar in gene expression. **(D)** BAL and nasal genes detected in both body sites were normalized to sequencing depth and modeled according to body site, age, and sex with clustering at the level of sample pair using an independent correlation matrix in generalized estimating equations. Model estimates for the relationship between body site and gene expression levels are plotted on the x-axis and FDR-adjusted p-values are plotted on the y-axis. Negative estimates indicate genes with higher expression in BAL (blue) and positive estimates indicate genes with higher expression in nasal swabs (pink). Example genes are shown in grey. **(E)** Normalized counts of differentially expressed genes are plotted in box-whisker plots where the box indicates the interquartile range and whiskers indicate observations within 1.5*IQR above or below the box. A violin plot is overlayed. Blue boxes represent BAL samples and pink boxes represent nasal samples. All comparisons are statistically significantly different at p_adjust_<0.05.

**Table 1:**
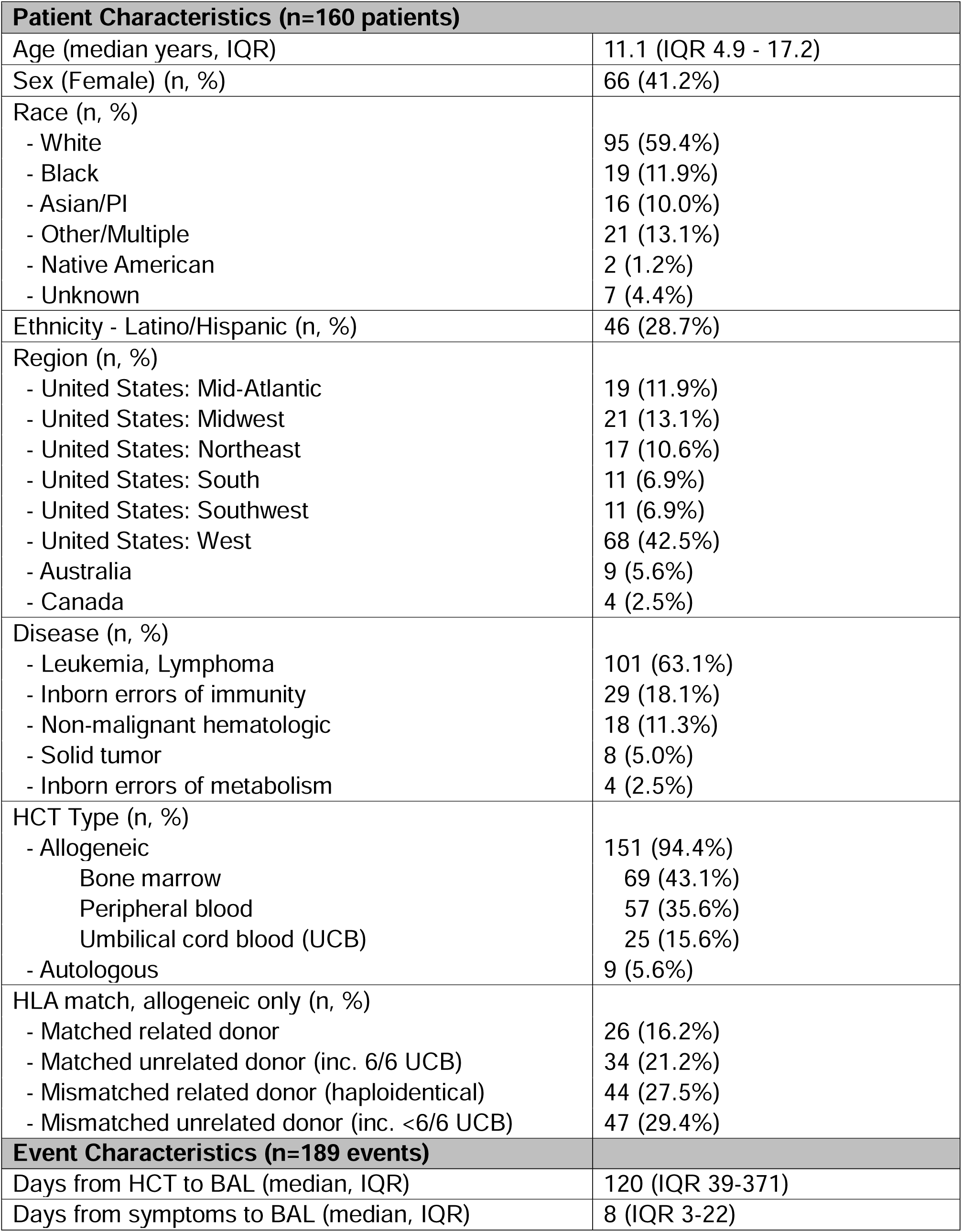
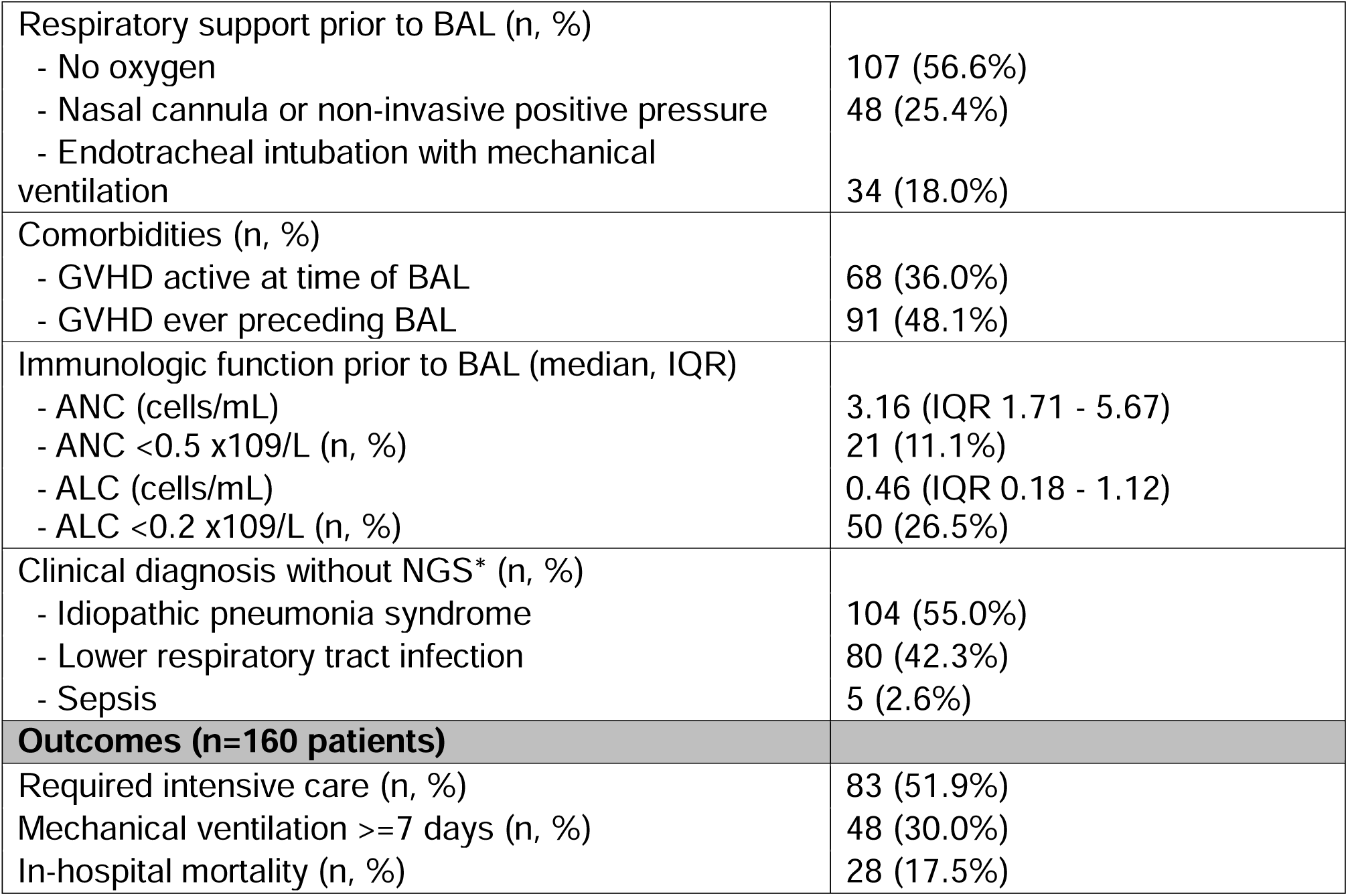
Patient Characteristics. **Legend**: Data for each patient (n=160) and each clinical event of BAL and nasal swab collection (n=189) are shown. Count data are described with numbers and percentages; distributions are described with median and interquartile range (IQR). * With reclassification based on mNGS results, 50 cases of IPS (26.5%) and 134 cases of LRTI (70.9%), see PMID 38783139 and processed data files for details.

| **Patient Characteristics (n=160 patients)** |  |
| --- | --- |
| Age (median years, IQR) | 11.1 (IQR 4.9 - 17.2) |
| Sex (Female) (n, %) | 66 (41.2%) |
| Race (n, %) |  |
| - White | 95 (59.4%) |
| - Black | 19 (11.9%) |
| - Asian/PI | 16 (10.0%) |
| - Other/Multiple | 21 (13.1%) |
| - Native American | 2 (1.2%) |
| - Unknown | 7 (4.4%) |
| Ethnicity - Latino/Hispanic (n, %) | 46 (28.7%) |
| Region (n, %) |  |
| - United States: Mid-Atlantic | 19 (11.9%) |
| - United States: Midwest | 21 (13.1%) |
| - United States: Northeast | 17 (10.6%) |
| - United States: South | 11 (6.9%) |
| - United States: Southwest | 11 (6.9%) |
| - United States: West | 68 (42.5%) |
| - Australia | 9 (5.6%) |
| - Canada | 4 (2.5%) |
| Disease (n, %) |  |
| - Leukemia, Lymphoma | 101 (63.1%) |
| - Inborn errors of immunity | 29 (18.1%) |
| - Non-malignant hematologic | 18 (11.3%) |
| - Solid tumor | 8 (5.0%) |
| - Inborn errors of metabolism | 4 (2.5%) |
| HCT Type (n, %) |  |
| - Allogeneic | 151 (94.4%) |
| Bone marrow | 69 (43.1%) |
| Peripheral blood | 57 (35.6%) |
| Umbilical cord blood (UCB) | 25 (15.6%) |
| - Autologous | 9 (5.6%) |
| HLA match, allogeneic only (n, %) |  |
| - Matched related donor | 26 (16.2%) |
| - Matched unrelated donor (inc. 6/6 UCB) | 34 (21.2%) |
| - Mismatched related donor (haploidentical) | 44 (27.5%) |
| - Mismatched unrelated donor (inc. <6/6 UCB) | 47 (29.4%) |
| **Event Characteristics (n=189 events)** |  |
| Days from HCT to BAL (median, IQR) | 120 (IQR 39-371) |
| Days from symptoms to BAL (median, IQR) | 8 (IQR 3-22) |
| Respiratory support prior to BAL (n, %) |  |
| - No oxygen | 107 (56.6%) |
| - Nasal cannula or non-invasive positive pressure | 48 (25.4%) |
| - Endotracheal intubation with mechanical ventilation | 34 (18.0%) |
| Comorbidities (n, %) |  |
| - GVHD active at time of BAL | 68 (36.0%) |
| - GVHD ever preceding BAL | 91 (48.1%) |
| Immunologic function prior to BAL (median, IQR) |  |
| - ANC (cells/mL) | 3.16 (IQR 1.71 - 5.67) |
| - ANC <0.5 x10<sup>9</sup>/L (n, %) | 21 (11.1%) |
| - ALC (cells/mL) | 0.46 (IQR 0.18 - 1.12) |
| - ALC <0.2 x10<sup>9</sup>/L (n, %) | 50 (26.5%) |
| Clinical diagnosis without NGS* (n, %) |  |
| - Idiopathic pneumonia syndrome | 104 (55.0%) |
| - Lower respiratory tract infection | 80 (42.3%) |
| - Sepsis | 5 (2.6%) |
| **Outcomes (n=160 patients)** |  |
| Required intensive care (n, %) | 83 (51.9%) |
| Mechanical ventilation ≥7 days (n, %) | 48 (30.0%) |
| In-hospital mortality (n, %) | 28 (17.5%) |

### Upper and lower airway gene expression

We detected expression of 15,133 protein-coding genes in nasal swabs (median 13,692, IQR 12,577-14,579) and 18,460 protein-coding genes in BALF (median 18,142, IQR 16,953-18,433). Paired nasal and BAL samples clustered separately in MDS space, indicating widespread transcriptional differences between upper and lower airways (**Figure 1C**). Of 15,114 genes detected in both body sites, 13,698 were differentially expressed by body site in models accounting for age, sex, repeat sampling events, and the paired nature of nasal and BAL sampling (FDR-adjusted p<0.05, **Data File 1, Figure 1D**). Examples of epithelial and immune genes highly enriched in the nose (CXCL8, MUC5AC, PIGR) and BALF (CD163, MARCO, FN1) are depicted in **Figure 1E**. Since both nasal swabs and BALF are composed of heterogeneous cellular populations, we contextualized bulk transcriptomes by deconvoluting 20 nasal and 28 BAL cell types (**Figure E1A)**. In comparing each nasal sample to its paired BAL, nasal samples generally showed a greater percentage of epithelial cell types (median difference +10.3%, IQR 2.5-22.7%), a lower percentage of myeloid cell types (median difference –8.5%, IQR –24.1% to –2.3%), and a similar percentage of lymphoid cell types (median difference 0.0%, IQR –3.1% to +3.2%), although exact cell types imputed within these categories varied (**Figure E1B**).

### Site-specific gene expression networks

We next considered that regardless of expression levels, the way gene networks were organized might differ between the upper and lower airways and could illustrate shared and unique functions. Therefore, we computed gene set enrichment scores to the 50 MSigDB Hallmark Gene Sets and tested for differential correlations by body site (**Data File 2, Figure 2A**). 130 gene set pairs demonstrated positive correlation within both nasal swabs and BALF; for example; expression of the *Hallmark Inflammatory Response* and *Hallmark Hypoxia* gene sets was positively correlated in both nasal swabs (ρ=0.70, p_adjusted_<0.001) and in BALF (ρ=0.76, p_adjusted_<0.001, **Figure 2B**), indicating a similar relationship in the upper and lower airways. In contrast, 57 gene set pairs demonstrated correlated expression in BALF (ρ≥0.5) but not in nasal swabs (ρ≤0.1), indicating differential gene regulation in the lower airways. The most common pathway in this list was *Hallmark Inflammatory Response*, which was correlated with 14 of 49 other gene sets in BAL but not in nasal swabs. For example, expression of the *Hallmark Inflammatory Response* and *Hallmark Reactive Oxygen Species* pathways were positively correlated in BALF (ρ=0.65, p_adjusted_<0.001) but not in nasal swabs (ρ=0.09, p_adjusted_=0.234, test of differences p_adjusted_<0.001).

**Figure 2.**
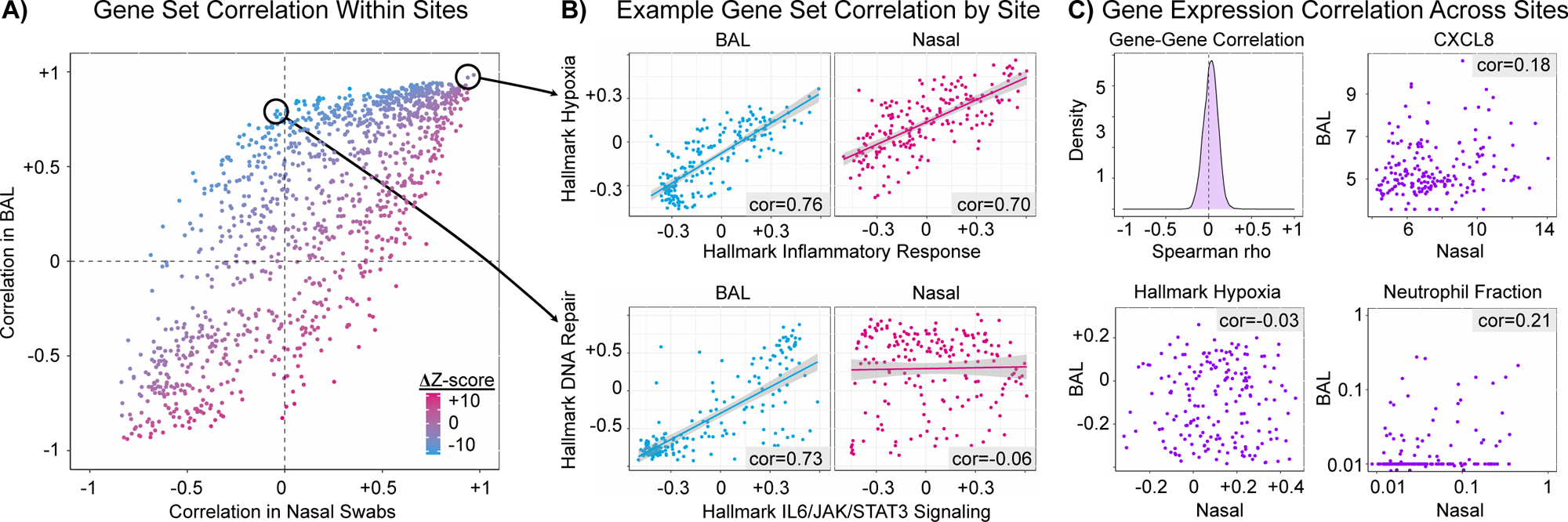
**Legend: (A)** Gene set enrichment scores to the 50 MSigDB Hallmark Gene Sets were calculated for BAL and nasal swabs together (*gsva*). Enrichment scores for each gene set pair were then correlated within BAL and within nasal swabs using Spearman rank-based correlation and plotted on the y– and x-axes, respectively. To contrast BAL and nasal correlations, correlation values were transformed to Z-scores (Fisher transformation) and the difference in Z-scores divided by the standard error of the difference is reported as ΔZ-score (dot shading, *ddcor*). Blue shaded dots indicate greater correlation in BAL than nasal; purple shaded dots indicate similar correlation in BAL and nasal; pink shaded dots indicate greater correlation in nasal than BAL. **(B)** Top: An example of similarly correlated gene sets in both BAL and nasal is depicted. Enrichment scores to Hallmark Inflammatory Response and Hallmark Hypoxia are positively correlated in both BAL and in nasal swabs. Bottom: An example of differently correlated gene sets in BAL and nasal swabs is depicted. Enrichment scores to Hallmark IL6/JAK/STAT3 signaling and Hallmark DNA Repair are positively correlated in BAL but not correlated in nasal swabs. **(C)** Top left: For each gene detected in both body sites (n=15,114), Spearman rank-based correlation was calculated for normalized gene expression between BAL and nasal swab pairs. A density distribution is plotted showing the distribution of Spearman rho values. Top right, Bottom left, Bottom right: Normalized expression of CXCL8, gene set enrichment score to the Hallmark Hypoxia Pathway, and imputed Neutrophil Fractions are plotted for BAL (y-axis) and nasal swabs (x-axis) with Spearman correlation listed in gray.

### Cross-site transcriptome correlation

Regardless of differences in expression levels or cellular compositions, correlation in expression between paired nasal and BAL samples could indicate shared regulation or common influences linking the upper and lower airways. However, nasal-BAL correlation for the 15,114 genes expressed in both tissues was minimal (median Spearman ρ=0.03, IQR –0.03 to 0.08; **Data File 3, Figure 2C**). Similarly, minimal correlations were observed between upper and lower airway expression of the 50 MSigDB Hallmark Gene Sets (median ρ=0.11, IQR 0.04-0.16, **Data File 4**) as well as between upper and lower airway cell types (median ρ=0.01, IQR –0.06 to +0.07, **Data File 5**). For example, BAL and nasal neutrophil fractions showed minimal correlation (ρ=0.212, p_adjusted_=0.131), as did BAL and nasal CD8+ T-cell fractions (ρ=0.042, p_adjusted_=0.860). ***Subgroup effect:*** We next hypothesized that the correlation between nasal and BAL gene expression would be higher in sicker patients due to heightened illness severity and systemic multi-organ dysfunction. When analyzing samples from just patients with the composite outcome of death or ≥7 days of mechanical ventilation, we detected stronger correlations between nasal and BAL expression of MSigDB Hallmark pathways such as IFNγ Response, IL6/JAK/STAT3 signaling, and TNF signaling via NFκB (p_adjusted_<0.05, **Data File 6**).

### Influence of immune reconstitution

We hypothesized that immune reconstitution status would be a primary driver of the differences in upper and lower airway gene expression observed between subjects. However, in models adjusted for age, sex, and day post-HCT, peripheral blood ANC was associated with expression levels of only 95 genes in nasal swabs and 16 genes in BALF (p_adjusted_<0.05, **Data Files 7-8**). Similarly, blood ANC was weakly associated with nasal neutrophil fraction (ρ=0.157, p=0.035) and was not associated with BAL neutrophil fraction (ρ=0.062, p=0.408). Similarly, peripheral blood ALC was not associated with expression levels of any genes in nasal swabs nor in BALF (p_adjusted_>0.05, **Data Files 9-10**). Blood ALC was weakly associated with nasal lymphoid fraction (ρ=0.169, p=0.023) and not associated with BAL lymphoid fraction (ρ=0.124 p=0.096). These data suggest that basic blood markers of immune status do not strongly explain the variation in BAL and nasal transcriptomes observed in this cohort.

### Upper and lower airway microbiome composition

The upper and lower respiratory tracts exist in a state of continuous exposure to myriad microbes, which exert reciprocal influence over epithelial and immune functions at the mucosal interface (27, 28). In this cohort, we detected RNA from a median 72 species (IQR 36-124) covering 34 genera (IQR 22-51) in nasal swabs and a median 76 species (IQR 27-145) covering 38 genera (IQR 17-73) in BALF, with similar α-diversity in both body sites (median nasal Simpson 0.61, IQR 0.29-0.75 vs. median BAL Simpson 0.57, IQR 0.27-0.78, Wilcoxon paired sign-rank p=0.721). However, as with transcriptomes, paired nasal and BAL microbiomes clustered separately in MDS space, indicating widespread taxonomic differences between upper and lower airways (**Figure 3A**). The top 10 most abundant nasal taxa included *Staphylococcus, Corynebacterium, and Streptococcus,* whereas the top 10 most abundant BAL taxa consisted of a variety of commensals (e.g. *Prevotella, Veillonella, Streptococcus),* potential pathogens (e.g. *Pseudomonas),* and low-virulence viruses (e.g. *Torquetenoviruses,* **Figure 3B**).

**Figure 3.**
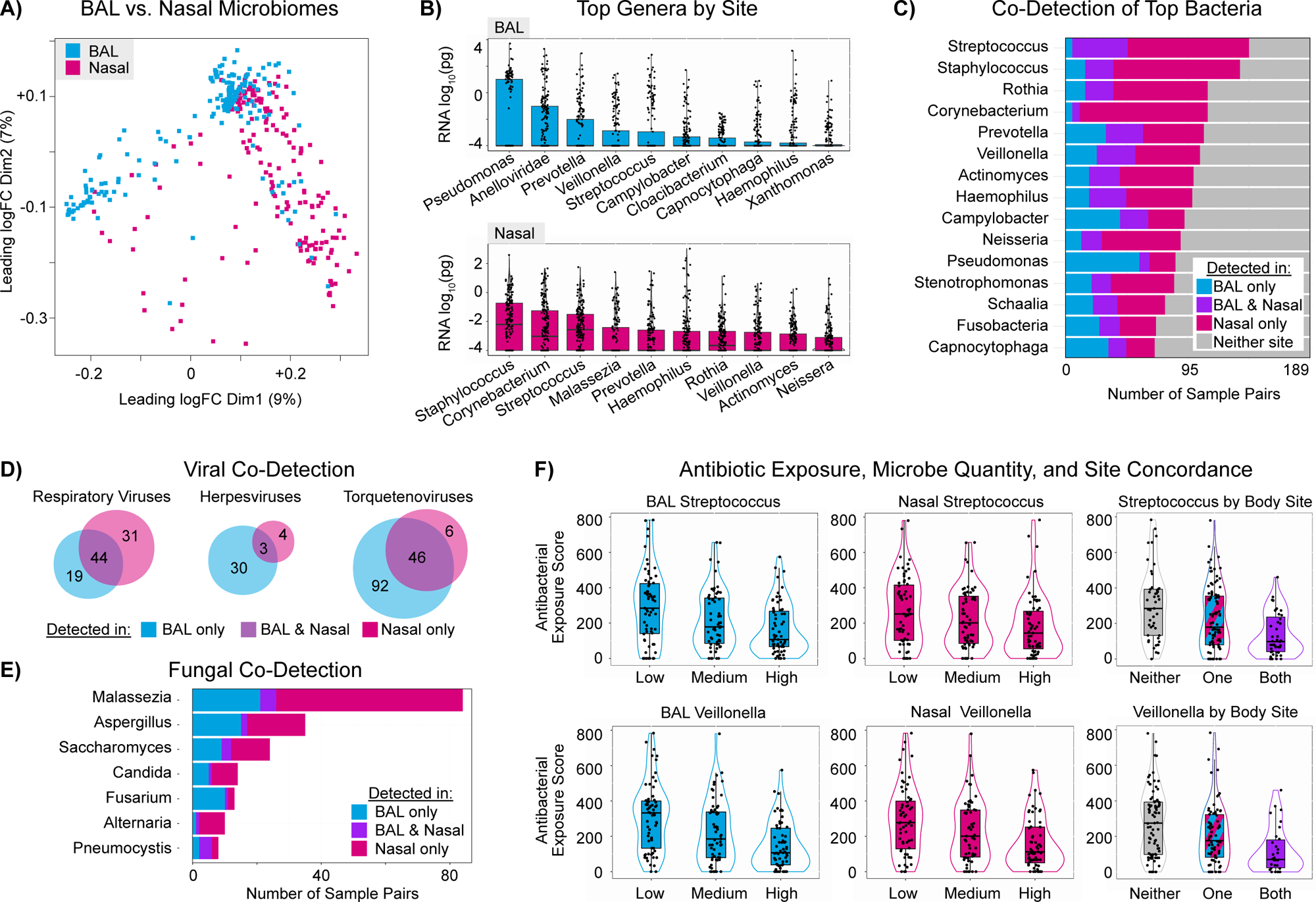
**Legend: (A)** BAL and nasal genus masses were filtered for taxa present in at least 5% of both sample types; sample distances were calculated and underwent multi-dimensional scaling (MDS), and samples were plotted according to the top two dimensions (x– and y-axes). Blue shading indicates BAL samples and pink shading indicates nasal samples. Samples closer in two-dimensional space are more similar in taxonomic composition. **(B)** Genus masses are plotted for BAL (blue) and nasal swabs (pink) using box-whisker plots arranged by descending order of the 75^th^ percentile value. **(C)** Every genus was labeled as detected in BAL only (blue), both BAL and nasal swab (purple), nasal swab only (pink), or neither body site (gray). The top 15 bacterial genera according to highest detection levels in the cohort are plotted with horizontal stacked bars. **(D)** Venn diagrams of number of respiratory viral genera, Herpesvirus genus, and Torquetenovirus genera detected in the cohort according to detection in BAL (blue), nasal swab (pink), or both (purple overlap). **(E)** The top 7 fungal genera according to highest detection levels in the cohort are plotted with horizontal stacked bars. **(F)** Antibacterial exposure score (AES) was calculated based on number of exposure days of each received agent, weighted by antibiotic broadness (see Supplemental Methods). The distribution of BAL Streptococcus RNA mass was split into tertiles (low, medium, high) and AES was plotted for each tertile (top left). This was repeated for Nasal *Streptococcus* (top middle). Sample-pairs were then categorized according to *Streptococcus* detection in neither, one, or both of the pair, and AES was again plotted (top right). This was then repeated for the *Veillonella* genus (bottom row).

We then tested for overlap between upper and lower microbiomes and found that most microbial genera were detected exclusively in either nasal or BAL samples rather than in both (**Data File 11-12, Figure 3C**). *Streptococcus pneumoniae* was the most frequently shared species but was detected in only 23% of sample pairs (43/189). Further, there was no correlation between upper and lower airway samples for aggregate microbiome traits such as richness, Shannon diversity, or total bacterial or fungal RNA (all p>0.05). However, total viral RNA was strongly correlated between upper and lower samples (ρ=0.54, p<0.001). Respiratory viruses detected in BAL samples were identified in 72% of paired nasal samples, whereas respiratory viruses detected in nasal samples were identified in 60% of paired BAL samples. In contrast, Herpesviruses and Torquetenoviruses were more commonly detected in BAL (**Figure 3D**). Fungal co-detection was rare, with only few cases of pathogens such as *Aspergillus*, *Candida*, and *Pneumocystis* being identified in paired samples (**Figure 3E**).

### Influence of antibiotics

We previously showed that BAL microbiomes are strongly influenced by the use of broad-spectrum antibiotics (9). In this cohort, antibiotic exposure preceded sample collection for 168/189 sample pairs, with a median antibacterial exposure score (AES) of 186 (IQR 79-349). In models adjusted for age, sex, and repeat sampling, AES was negatively associated with both nasal and BAL quantity of commensal bacteria such as *Haemophilus*, *Streptococcus*, *Veillonella*, and *Gemella* (p_adjusted_<0.05, **Data Files 13-14, Figure 3E**). We then questioned whether the degree of upper-lower microbiome discordance was related to antibiotic exposure and resultant commensal depletion. Here we found that greater AES was associated with loss of upper-lower airways taxa co-detection for commensal taxa in particular (**Data File 15**). For example, AES was lowest for patients with taxa such as *Streptococcus* or *Veillonella* detected in both upper and lower samples, moderate for patients with detection in one body site, and highest for patients without detection in either body site (p_adjusted_<0.05, **Figure 3F**).

### Integrating Human Gene Expression and the Microbiome Across Body Sites

Having described the microbiome and transcriptomes separately, we next aimed to identify microbiome-transcriptome relationships both consistent and unique across the upper and lower airways.

### Site-Specific Relationship with Bacteria

In nasal swabs, total bacterial RNA was associated with expression of 710 genes whereas in BAL, total bacterial RNA was associated with expression of 10,394 genes (p_adjusted_<0.05, **Figure 4A**). In both body sites, greater total bacterial RNA was positively associated with expression of 248 genes showing enrichment for a variety of inflammatory pathways. In nasal swabs but not BAL, total bacterial RNA was positively associated with an additional 352 genes also related to inflammatory pathways (**Figure 4B**). In contrast, in BAL but not nasal swabs, greater bacterial RNA was positively associated with expression of 9,265 genes related to a variety of epithelial pathways such as ion transport, cilium activity, and collagen deposition. Further, BAL bacterial RNA was negatively associated with expression of 850 genes related to myeloid cell function including IFNγ response and Fcγ-mediated phagocytosis. These findings were confirmed on statistical interaction testing (**Data Files 16-18**).

**Figure 4.**
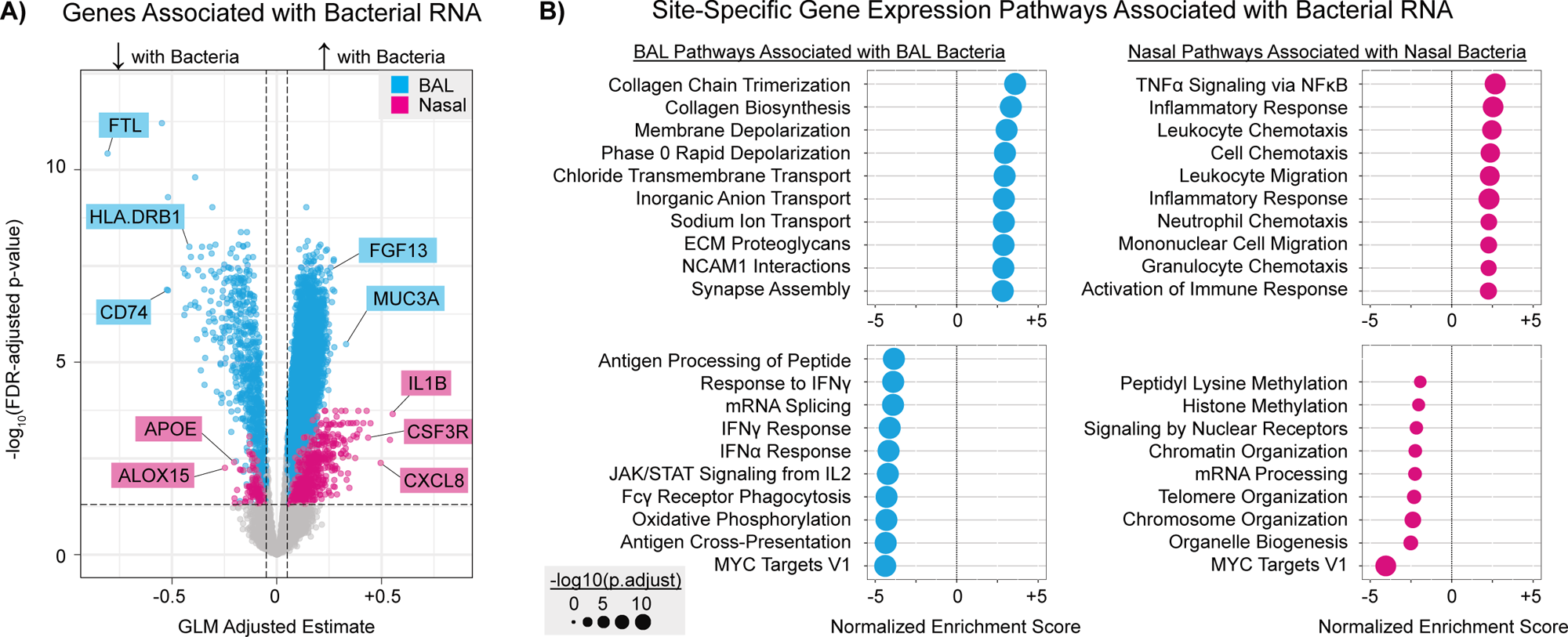
**Legend: (A)** Normalized gene counts were modeled according to log_10_-transformed total bacterial RNA with adjustment for age and sex and clustering at the level of sample pair using an independent correlation matrix in generalized estimating equations. Model estimates for the relationship between bacterial RNA and gene expression levels are plotted on the x-axis and FDR-adjusted p-values are plotted on the y-axis. Negative estimates indicate genes with lower expression with increasing bacterial RNA and positive estimates indicate genes with higher expression with increasing bacterial RNA. Blue dots indicate relationships in BAL and pink dots indicate relationships in nasal swabs. Example genes are shown in boxes for the respective color. **(B)** BAL genes whose expression levels were associated with quantity of BAL bacterial RNA underwent gene set enrichment analysis (*gsea*, *clusterProfiler*) to the Reactome, GO:BP, and Hallmark gene set repositories. The top 10 enriched gene sets according to highest and lowest normalized enrichment score (NES) are plotted, with dot size indicating FDR-adjusted p-value. This was repeated for nasal genes associated with quantity of nasal bacterial RNA (pink).

### Site-Specific Relationship with Viruses

In nasal swabs, total viral RNA was associated with expression of 25 genes with enrichment for IFNγ signaling. In contrast, in BALs, total viral RNA was associated with expression of 10,734 genes showing positive enrichment for epithelial-mesenchymal pathways such as collagen chain trimerization and anchoring fibril formation and negative enrichment for a variety of antigen-presenting cell processes such as phagocytosis, macrophage cytokine production, and detoxification of ROS (**Data Files 19-20**).

### Site-Specific Relationship with Fungi

In nasal swabs, total fungal RNA was not associated with expression of any genes after accounting for age, sex, and repeat sampling. In contrast, in BAL, total fungal RNA was associated with expression of 1,197 genes. Pathway enrichment indicated increased epithelial pathways such as keratinization and cornification and decreased T-cell-mediated immunity (**Data Files 21-22**).

### Airway Metatranscriptome and Clinical Outcomes

The composite outcome of in-hospital mortality or ≥7 days of mechanical ventilation occurred in 30% of patients (48/160). However, nasal swabs from these patients differentially expressed only 4 genes whereas BALs differentially expressed 5,676 genes (p_adjusted_<0.05, **Data Files 23-24, Figure 5A**). Pathway analysis indicated (1) increased BAL expression of epithelial-mesenchymal pathways such as collagen biosynthesis, smooth muscle contraction, and laminin interactions; and (2) decreased BAL expression of a variety of immune pathways such as granulocyte activation, IFNα response, and antigen processing and presentation (**Figure 5B**). No nasal microbiome features were associated with the composite outcome, whereas BAL microbiomes showed a lower quantity of commensal genera such as *Prevotella*, *Streptococcus*, and *Veillonella* (p_adjusted_<0.05, **Data Files 25-26**).

**Figure 5.**
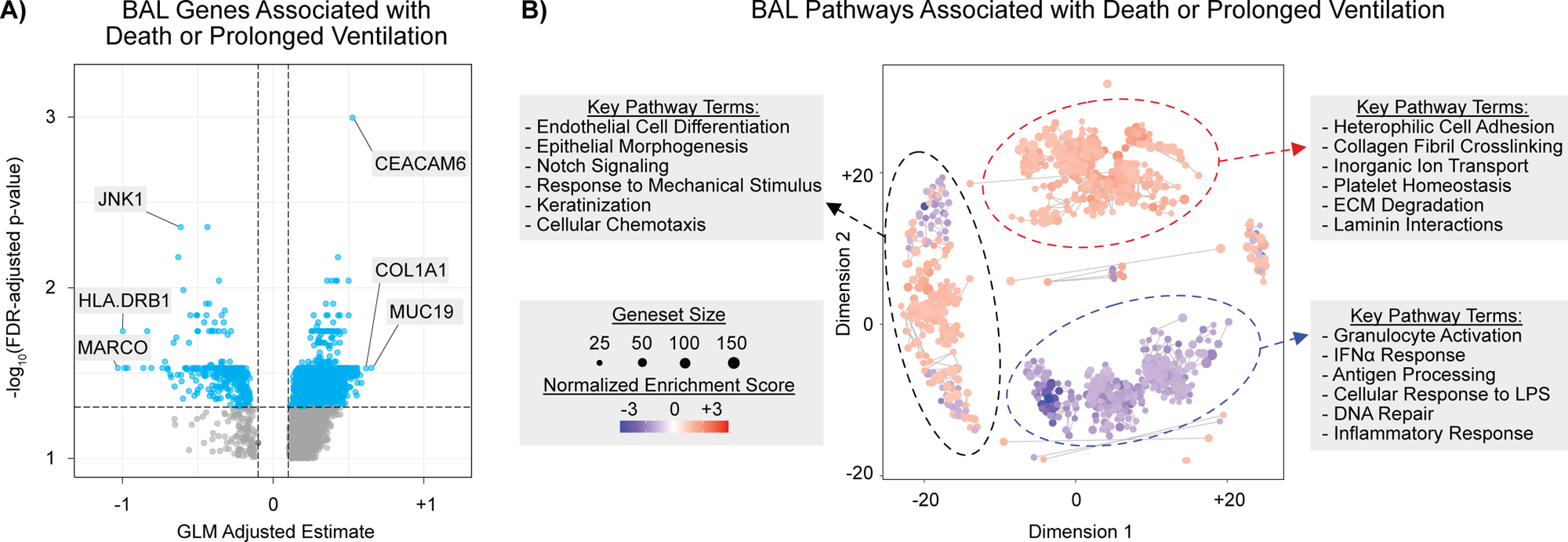
**Legend: (A)** Normalized gene counts were modeled according to the composite outcome of death or ≥7 days of mechanical ventilation with adjustment for age and sex and clustering at the level of sample pair using an independent correlation matrix in generalized estimating equations. Model estimates for the relationship between the outcome and gene expression levels are plotted on the x-axis and FDR-adjusted p-values are plotted on the y-axis. Negative estimates indicate genes with lower expression in patients with poor outcomes and positive estimates indicate genes with higher expression in patients with poor outcomes. Blue dots indicate relationships in BAL. Example genes are shown in boxes for the respective color. **(B)** 5,676 BAL genes associated the composite outcome underwent gene set enrichment analysis (*gsea*, *clusterProfiler*) to the Reactome, GO:BP, and Hallmark gene set repositories, yielding 870 enriched gene sets. Given the large number of enriched gene sets, gene sets were then tested for similarity based on shared gene lists (*clusterProfiler*); pairwise similarity scores underwent multi-dimensional scaling (MDS) and were plotted in 2-dimensional space. Each dot represents a gene set with dot size indicating number of genes in the gene set, and dot shading indicating increased (red) or decreased (blue) enrichment in patients with the composite outcome. Genesets closer in two-dimensional space are more similar. In grey, examples of key terms from gene set “clusters” are listed.

Finally, to validate this approach, we used an unsupervised machine learning algorithm to reduce the dimensionality of the paired upper and lower airway gene expression and microbiome measurements. Clustering on the resultant 15 latent dimensions identified 2 distinct patient groups, with group 1 containing nearly 80% of the patients with the composite outcome (38/104 vs 10/56, p=0.023, **Figure E2**). Both groups had similar demographics, transplant characteristics, and peripheral blood ANC and ALC (**Table E1**). In models adjusted for age, sex, and repeat sampling, the two groups differed significantly in BAL gene expression (n=11,442 DEGs) but not in nasal gene expression (n=0 DEGs), again confirming the lack of prognostic information from nasal swabs in this cohort (**Data Files 27-28**).

## DISCUSSION

In this study, we examined 189 paired nasal swabs and BAL samples from 160 pediatric patients with post-HCT lung injury. We hypothesized that upper and lower respiratory samples would show overlap in gene expression and microbiome features, but instead found minimal cross-site correlation, highlighting the unique biology of the alveolar space and indicating compartmentalization of lung injury response. Further evidence for compartmentalization of lung injury response in this population includes site-specific microbiome-transcriptome interactions and a major difference in clinical prognosis gleaned from BALF data relative to data from nasal swabs. Taken together, these results indicate that nasal sampling does not adequately reflect bronchoalveolar biology in this population and bronchoscopy ought to remain the priority for our field at this time.

A primary objective of this study was to determine whether nasal sampling could suitably approximate alveolar biology and microbiology in the post-HCT setting. Prior work has shown correlations between nasal and tracheobronchial transcriptomes in airway diseases such as asthma, COPD, bronchiolitis, and cystic fibrosis, as well as in healthy and smoking populations (29–39). In contrast, this study examined nasal-alveolar sample pairs in post-HCT patients with predominantly parenchymal disease, a population rarely represented in the literature. The absence of upper-lower airway transcriptome and microbiome concordance in this cohort likely reflects both the distinct sampling strategy (nasal vs. BAL) and the unique biology in the post-HCT setting. Whether different biologic markers such as proteomics or methylation assays might show stronger pan-respiratory correlation remains an ongoing question (40). In addition, it remains possible that more homogeneous subsets of post-HCT patients, such as those with bronchiolitis obliterans or similar respiratory viruses, may show greater upper-lower airway transcriptome correlation than what we observed here (41). Of note, we previously reported that BAL and paired blood samples also shared minimal transcriptomic overlap, further highlighting the compartmentalization observed in the post-HCT lungs (42).

A secondary objective of this study was to emphasize post-HCT pathobiology unique to the lungs. First, BAL samples were typically composed of a greater fraction of myeloid cells and fewer epithelial cells than nasal samples, which may be related to differences in sampling technique (lavage vs. brush). Second, BAL expression networks were organized differently. For example, in the lung—but not the nose—Hallmark Inflammatory Response was tightly associated with Reactive Oxygen Species, Cholesterol Homeostasis, and DNA Repair, indicating site-specific immunologic mechanisms linking inflammation and immunometabolism. Third, BAL microbiomes harbored fewer commensal taxa and more DNA viruses such as *Herpesviruses* and *Torquetenoviruses,* which may be due local differences in factors affecting microbial immigration, elimination, and in the cases of viruses, re-activation (15, 28, 43, 44). Fourth, the relationship between microbes and respiratory gene expression differed by site. For example, nasal bacteria were associated with nasal expression of inflammatory genes, whereas BAL bacteria were associated with a lack of local immune signaling. This may highlight the unique role of the nose as a first point of defense to the outside world in contrast to the protected local environment of the respiratory zone. Further, the quantity of bacterial, viral, and fungal RNA in the BAL were each associated with major epithelial programming shifts related to injury response, keratinization, and collagen deposition. Indeed, chronic pulmonary viral and fungal exposure has been linked to airway fibrosis in post-HCT bronchiolitis obliterans as well as in non-HCT disease such as IPF (45–48). Taken together, these findings highlight the unique epithelial-immune-microbial interactions in the post-HCT lung.

From a clinical standpoint, nasal samples are unlikely to adequately represent the bronchoalveolar environment, but could theoretically still have value as biomarkers in outcome prediction (49, 50). However, our data identified minimal prognostic information in nasal swabs for the composite outcome of death or prolonged mechanical ventilation, thus further questioning their utility in risk stratification. As such, ongoing identification of minimally-invasive tools for screening, diagnosis, and disease monitoring remains a barrier in the field of post-HCT lung injury.

Strengths of our study include the paired nature of upper and lower airway sampling, the large multicenter cohort, and the use of paired microbiome and gene expression platforms. Limitations include the use of bulk sequencing rather than single-cell tools and the heterogeneous nature of post-HCT lung toxicities included in this cohort.

In conclusion, our study highlights the compartmentalization of the alveolar space in pediatric HCT patients with a broad range of post-HCT lung toxicities. These data illuminate unique alveolar epithelial-immune-microbial interactions and emphasize the ongoing need for minimally-invasive diagnostics in this population.

## Data Availability

Raw sequencing files and instructions to request download are available under controlled access on NIH dbGaP: https://www.ncbi.nlm.nih.gov/projects/gap/cgi-bin/study.cgi?study_id=phs001684.v3.p1. Individual-level data are available indefinitely. Code and processed anonymized individual-level data files are available on GitHub: https://github.com/zinterm/pedBMT_BALseq. Results of statistical analyses are available in Supplemental Data Files.

## ACKNOWLEDGEMENTS

The authors thank Dr. Joseph L. DeRisi for his scientific guidance and manuscript review, as well as the children and families who participated in this study.

## Artificial Intelligence

The AI tool ChatGPT 5.0 was used to assist with RStudio coding syntax. All AI-provided coding suggestions were manually checked for accuracy.

## Author contributions

Conceptualization: MSZ, CCD, EE Methodology: MSZ, CCD, EE Investigation: all authors Visualization: MSZ, EE Funding acquisition: MSZ, CCD, EE Project administration: MSZ, CCD, EE Supervision: MSZ, CCD Writing – original draft: MSZ, CCD, EE Writing – review & editing: all authors

## Online Data Supplement

This article has an online data supplement, which is accessible from this issue’s table of content online at www.atsjournals.org

**Figure E1.**
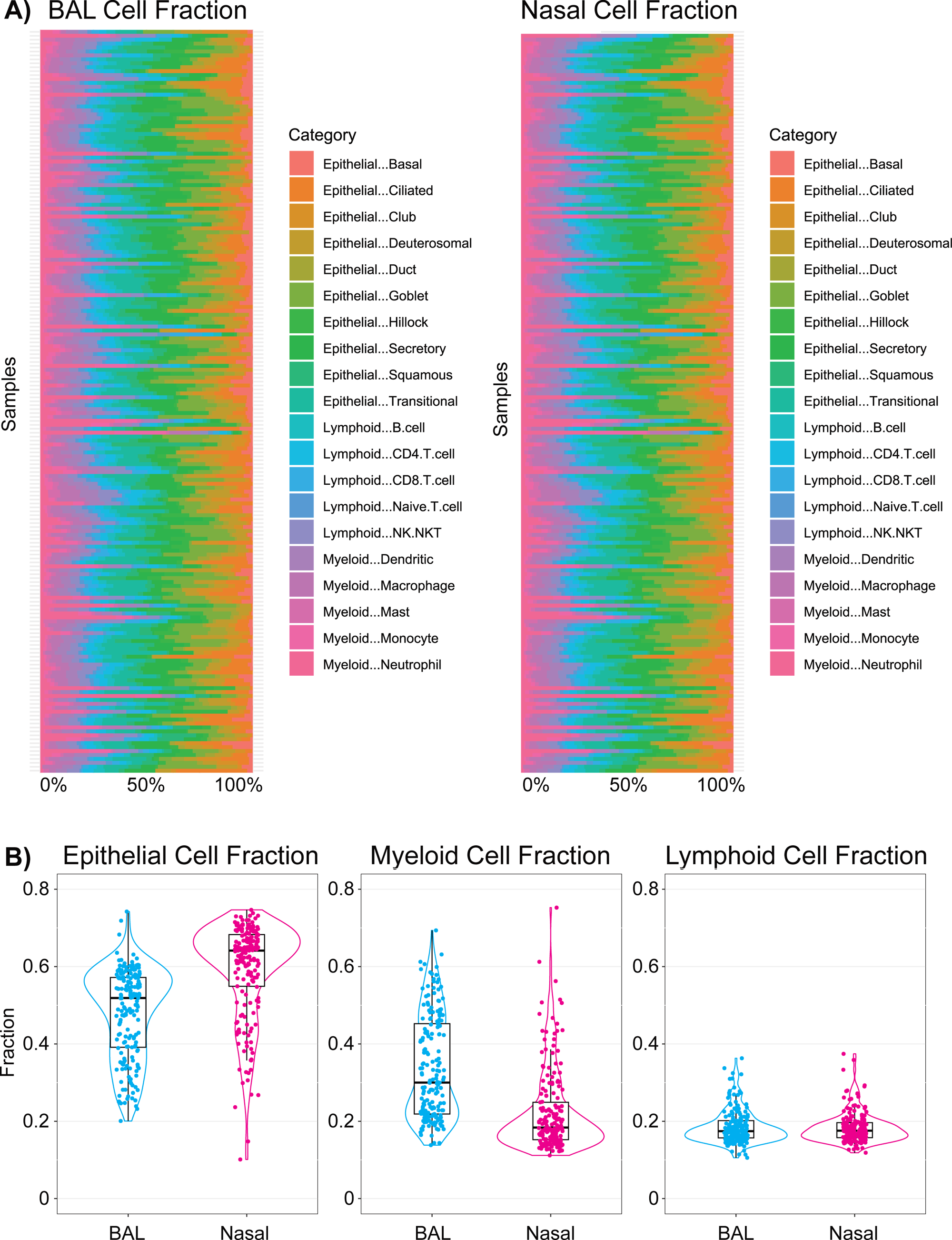
**Legend: (A)** Cell fractions were imputed using CIBERSORTx and the LungMAP reference atlas for BAL (Guo et al, Nat Comm 2023) and the Yoshida et al reference atlas for nasal swabs (Nature, 2022). Each sample is listed as a row; horizontal stacked bars indicate imputed cell fractions adding up to 100%. **(B)** Cell fractions were summed at the level of epithelial, myeloid, and lymphoid populations and summed fractions are plotted for BAL (blue) and nasal (pink) using box-whisker plots with violins.

**Figure E2.**
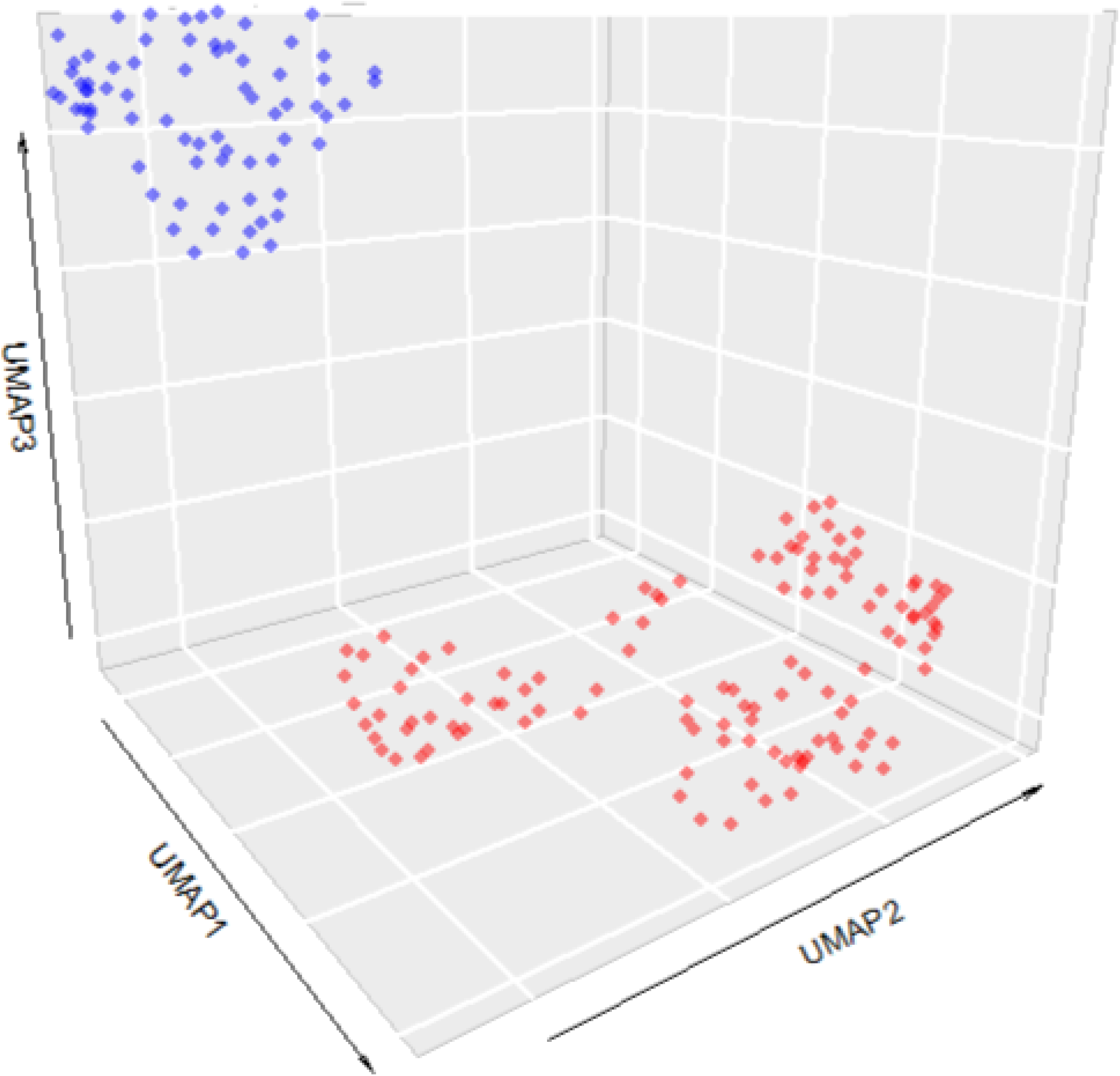
**Legend**: Microbiome and gene expression matrices for BAL and nasal swabs, each linked by patient, underwent multi-omics factor analysis (*mofa*) with dimensionality of the resultant 15 latent factors then reduced using umap. Each sampling event (n=189 events) is plotted in 3-dimensional space, with clear separation of 2 clusters shown in red (cluster 1) and blue (cluster 2).

