## Supplemental Material for "Upper and Lower Respiratory Tract Compartmentalization in Pediatric Stem Cell Transplantation"

***Patients:*** Details of patient enrollment are previously described (PMID 38783139). Participating pediatric centers screened all HCT patients preparing to undergo clinically-indicated bronchoscopic BAL for diagnostic assessment of pulmonary disease of any suspected type, including infectious or non-infectious etiologies (NCT02926612). Typically patients presented with respiratory symptoms (e.g. cough, dyspnea, work of breathing) and/or chest imaging findings (e.g. ground glass nodules, peribronchial thickening, opacities), and/or decline in pulmonary function testing concerning for occult infection or alloreactive process (e.g. GVHD). Frequently, multiple etiologies were on the differential diagnosis, hence necessitating the bronchoscopy. The study did not dictate if and when bronchoscopies were performed. Of note, patients with clear reasons for post-HCT lung injury (e.g. blood culture positive sepsis, nasal PCR positive for influenza, etc) may be under-represented in this cohort since their treating teams may have felt bronchoscopy was unnecessary for diagnostic confirmation. Patients or their guardians were approached for consent prior to specimen collection with local IRB approval at each site (UCSF IRB #14-13546, #16-18908) in accordance with the Declaration of Helsinki.

***Samples:*** Details of sample procurement are previously described (PMID 38783139). BAL was performed by pediatric pulmonologists trained in fiberoptic bronchoscopy using institutional protocols. Typically aliquots of BAL were pooled and not separated for analysis. Target area of the lung for bronchoscopy was at the discretion of the clinical pulmonologist. Volume lavaged and volume returned were not documented. After aliquoting BALF for clinical testing, excess lavage was aliquoted into sterile cryovials. Anterior nasal swabs were obtained during the BAL procedure using sterile Floq-swabs placed in conical tubes without media. Both BALF and nasal swabs were placed immediately on dry ice and stored at -70ºC until processing. Typically sample aliquoting occurred in the operating room or ICU rather than in a central processing lab, and thus time from sample collection to dry ice was <15 minutes.

***Clinical Metadata:*** All clinical metadata were obtained from chart review by trained research coordinators at each site. As previously reported, all doses of antimicrobials administered in the 7 days before BAL were documented. The Antibacterial Exposure Score (AES) was calculated by summing the days of exposure to each antibacterial agent weighted with an agent-specific broadness score ranging from 4 to 49.75 (for example, ampicillin 13.50, meropenem 41.50) using the weighting scores derived by Madaras-Kelly et al, 2014. Daily dosages were not collected.

***Data Generation:*** Details of BAL processing are previously described (PMID 38783139) and were optimized in prior publications (PMIDs 35263147, 33512420, 30239621, 30992055, 30629604). Briefly, 200uL of thawed, unfractionated BALF underwent mechanical homogenization in RNA-preserving media followed by column-based RNA extraction (Zymo) and non-directional library preparation (NEB Ultra II). Specific RNA depletion or enrichment steps were not used.

For this study, 2 nasal swabs per patient collected in a conical 15mL tube were combined with 1mL RNA-preserving media (Zymo DNA/RNA Shield 2x) and the tube was vortexed on a tabletop instrument for 15 seconds to release mucous. Subsequently, swabs were cut with sterile scissors and placed into a 2mL tube containing 0.5mm glass beads (Tough Micro-Organism Lysing Mix 2 mL Tubes Nuclease & Microbial DNA Free, Omni International, SKU:19-622D400uL). Then the 1mL RNA-preserving media was add to the 2mL bead-bashing tube and sealed. The tube then underwent mechanical homogenization for 25 seconds at 30Hz followed by rest on ice for 60 seconds (TissueLyserII, Qiagen). This was repeated for 5 total cycles, after which the tube was centrifuged for 10 minutes at 16,000 rpm at 4°C. 200uL of the resultant solution was used for RNA extraction. RNA extraction was optimized for a Beckman Coulter Biomek NX^P^ instrument using the Zymo Quick-RNA Magbead kit, Cat. No. R2133, with a DNase I step per the manufacturer’s protocol. Briefly, 200uL sample was combined with 600uL RNA Lysis Buffer, mixed for 5 minutes, then that 800uL solution was combined with 800uL of 100% EtOH and mixed for another 5 minutes. Next, 30uL MagBinding Beads were added and mixed for 20 minutes. Samples were transferred to a magnetic plate to facilitate removing the supernatant while retaining beads in the tube. Beads were then washed with 500uL RNA Wash 1, supernatant was removed via magnetic bead binding, and washed again with 500uL RNA Wash 2, followed by 2 additional washes with 500uL 100% EtOH. Samples were then incubated with 50uL DNase mix and mixed gently for 10 minutes, followed by additional washes with 500uL Prep Buffer and 500uL 100% EtOH (EtOH wash performed twice). Finally, samples were eluted in 45uL sterile water and this solution of purified RNA was used for library preparation in the same manner as the BAL RNA.

As previously described, library preparation utilized a miniaturized version of New England Biolabs UltraII Non-Directional RNA Library Prep Kit (protocol published previously: dx.doi.org/10.17504/protocols.io.tcaeise). Reagents were dispensed using the Echo 525 (Labcyte) and underwent Ampure-XP bead cleaning on a Beckman Coulter Biomek NX^P^ instrument. Libraries underwent 19 cycles of polymerase chain reaction (PCR) amplification, size selection to a target 300 to 700 nucleotides (nt), and were pooled to facilitate approximately even depth of sequencing. Twenty-five picograms (pg) of External RNA Controls Consortium (ERCC) pooled standards were spiked-in to each sample after RNA extraction and before library preparation to serve as internal positive controls (Thermo Fisher Scientific Cat. No 4456740). In addition, to identify contamination in laboratory reagents and the laboratory environment, each batch contained 2 samples of nasal swabs that were unopened/unused prior to RNA extraction, as well as nasal swabs mixed with 200 µL HeLa cells taken from a laboratory stock and processed identically to patient samples, in order to account for laboratory- and reagent-introduced contamination. Samples were first sequenced at shallow depth on an Illumina iSeq instrument and subsequently pooled across lanes of an Illumina NovaSeq X instrument and sequenced to a target depth of 40 million read-pairs with sequencing read length of 125 nt. Resultant fastq files were processed using CZID pipeline version 7.1, which is described in detail on czid.org (PMID 33057676). Briefly, this protocol involves serial rounds of human read alignment (STAR to hg38, Bowtie 2, STAR again, GSNAP) as well as Illumina adapter removal (Trimmomatic), quality filtering (PriceSeq package) and Lempel–Ziv–Welch complexity filtering. After hg38 aligning transcripts were identified and set aside, remaining human-subtracted files underwent alignment to the NCBI nt/nr database using GSNAP with a minimum alignment length greater than 36. Duplicate reads were added back in and taxa counts were generated with associated metrics of percentage identity, contig length and e-value to the nearest NCBI hit. To reduce spurious associations due to ambiguous alignments, taxa were excluded if they (1) aligned to archaea or uncultured microorganisms, (2) had 6 or fewer total reads, (3) had less than 100 nt alignment length, or (4) had less than 80%, 90% or 95% nucleotide percentage

identity for viruses, eukaryotes and bacteria, respectively. In addition, samples with low biomass (less than 100 pg) were further filtered to keep only taxa with 10 or more transcripts forming a contig of 250 nt or more with 80% or more percentage identity to the nearest NCBI hit. These criteria are the same as used for BAL samples.

***Analysis:*** Code and processed anonymized individual-level data files are available on GitHub: <https://github.com/zinterm/pedBMT_BALseq>. All code was performed in RStudio 2025.05.1 using R 4.4.0. Clinical data were summarized as counts with percentages or medians with interquartile ranges. BAL and nasal transcriptomes were compared graphically using MDS plots (*edgeR*) and normalized counts (produced using *variancestabilizingtransformation* in *DESeq2*) were modeled against body site, peripheral blood immune cell counts, BAL taxonomic domains, and clinical outcomes using generalized estimating equations accounting for age, sex, and repeat samples from the same patient (*geeglm* functions). A Gaussian distribution was used and an independence correlation structure was used for repeat observations of the same patient. Differentially expressed genes underwent pathway enrichment using MSigDB, Reactome, and GOBP datasets using over-representation analysis (ORA) or gene set enrichment analysis (GSEA) where appropriate (functions in the *ClusterProfiler* package). Gene set enrichment scores were calculated using the *gsva* package. Differential correlation of gene sets by body site was tested using the *DGCA* package. BAL and nasal transcriptomes were compared within patients by testing for Spearman rank-based correlation of normalized gene counts (*cor.test* and *rcorr*). BAL and nasal cell types were estimated using CIBERSORTx with recent reference cell atlases. Analyses were repeated at the microbial level by substituting human gene expression for taxonomic counts. All analyses involving more than 10 comparisons underwent FDR-adjustment of p-values. Finally, BAL and nasal gene expression and taxonomic data were modeled together using machine learning followed by dimensionality reduction (*MOFA*) with standard parameters. Resultant 15 latent factors underwent dimensionality reduction with *umap* (_neighbors = 70, min_dist=0, n_components=3, negative_sample_rate=70) followed by k-means clutsering (*kmeans*) with the ideal number of clusters determined by the Elbow and Silhouette methods (*factoextra*). Data were visualized using volcano plots (*EnhancedVolcano*), box-whisker violin plots (*ggplot2*), heatmaps (*pheatmap*), and gene set enrichment plots (*ClusterProfiler*).

**Table E1: Patient Characteristics**

| **Patient Characteristics (n=160 patients)** |  | **Cluster 2 (red)** | **Cluster 1 (blue)** | **Sig.** |
| --- | --- | --- | --- | --- |
| Age (median years, IQR) | 11.1 (IQR 5.2 – 16.2) | | 11.3 (4.1-17.5) | P=0.994 |
| Sex (Female) (n, %) | 44 (42.3%) | | 22 (39.3) | P=0.840 |
| Race (n, %) |  | |  | P=0.953 |
| - White | 61 (58.7%) | | 34 (60.7%) |  |
| - Black | 14 (13.5%) | | 5 ( 8.9%) |  |
| - Asian/PI | 10 ( 9.6%) | | 6 (10.7%) |  |
| - Other/Multiple | 13 (12.5%) | | 8 (14.3%) |  |
| - Native American | 1 ( 1.0%) | | 1 ( 1.8%) |  |
| - Unknown | 5 ( 4.8%) | | 2 ( 3.6%) |  |
| Ethnicity - Latino/Hispanic (n, %) | 28 (26.9%) | | 18 (32.1%) | P=0.608 |
| Region (n, %) |  | |  | P=0.725 |
| - United States: Mid-Atlantic | 13 (12.5%) | | 6 (10.7%) |  |
| - United States: Midwest | 12 (11.5%) | | 9 (16.1%) |  |
| - United States: Northeast | 12 (11.5%) | | 5 ( 8.9%) |  |
| - United States: South | 6 ( 5.8%) | | 5 ( 8.9%) |  |
| - United States: Southwest | 8 ( 7.7%) | | 3 ( 5.4%) |  |
| - United States: West | 42 (40.4%) | | 26 (46.4%) |  |
| - Australia | 8 ( 7.7%) | | 1 ( 1.8%) |  |
| - Canada | 3 ( 2.9%) | | 1 ( 1.8%) |  |
| Disease (n, %) |  | |  | P=0.547 |
| - Leukemia, Lymphoma | 65 (62.5%) | | 36 (64.3%) |  |
| - Inborn errors of immunity | 17 (16.3% | | 12 (21.4%) |  |
| - Non-malignant hematologic | 12 (11.5%) | | 6 (10.7%) |  |
| - Solid tumor | 6 ( 5.8%) | | 2 ( 3.6%) |  |
| - Inborn errors of metabolism | 4 ( 3.8%) | | 0 ( 0.0%) |  |
| HCT Type (n, %) |  | |  | P=1.00 |
| - Allogeneic | 98 (94.2%) | | 53 (94.6%) |  |
| Bone marrow | 50 (48.1%) | | 19 (33.9%) |  |
| Peripheral blood | 36 (34.6%) | | 21 (37.5%) |  |
| Umbilical cord blood (UCB) | 12 (11.5%) | | 13 (23.2%) |  |
| - Autologous | 6 ( 5.8%) | | 3 ( 5.4%) |  |
| HLA match, allogeneic only (n, %) |  | |  | P=0.847 |
| - Matched related donor | 18 (17.3%) | | 8 (14.3%) |  |
| - Matched unrelated donor (inc. 6/6 UCB) | 24 (23.1%) | | 10 (17.9%) |  |
| - Mismatched related donor (haploidentical) | 26 (25.0%) | | 18 (32.1%) |  |
| - Mismatched unrelated donor (inc. <6/6 UCB) | 30 (28.8%) | | 17 (30.4%) |  |
| **Event Characteristics (n=189 events)** |  | |  |  |
| Days from HCT to BAL (median, IQR) | 119 (IQR 42-329) | | 138 (IQR 37-442) | P=0.761 |
| Days from symptoms to BAL (median, IQR) | 8 (IQR 3-26) | | 7 (IQR 2-14) | P=0.311 |
| Respiratory support prior to BAL (n, %) |  | |  | P=0.142 |
| - No oxygen | 59 (50.4%) | | 48 (66.7%) |  |
| - Nasal cannula or non-invasive positive pressure | 34 (29.1%) | | 14 (19.4%) |  |
| - Endotracheal intubation with mechanical ventilation | 24 (20.5%) | | 10 (13.9%) |  |
| Comorbidities (n, %) |  | |  |  |
| - GVHD active at time of BAL | 46 (39.3%) | | 22 (30.6%) | P=0.466 |
| - GVHD ever preceding BAL | 61 (52.1%) | | 30 (41.7%) | P=0.374 |
| Immunologic function prior to BAL (median, IQR) |  | |  |  |
| - ANC (cells/mL) | 3.22 (1.74-6.11) | | 2.69 (1.50-5.03) | P=0.231 |
| - ALC (cells/mL) | 0.42 (0.17-1.16) | | 0.54 (0.20-0.99) | P=0.619 |
| Clinical diagnosis without NGS* (n, %) |  | |  |  |
| - Idiopathic pneumonia syndrome | 55 (47.0%) | | 49 (68.1%) | P=0.001 |
| - Lower respiratory tract infection | 61 (52.1%) | | 19 (26.4%) |  |
| - Sepsis | 1 ( 0.9%) | | 4 ( 5.6%) |  |
| **Outcomes (n=160 patients)** |  | |  |  |
| Required intensive care (n, %) | 54 (51.9%) | | 29 (51.8%) | P=1.00 |
| Mechanical ventilation >=7 days (n, %) | 38 (36.5%) | | 10 (17.9%) | P=0.023 |
| In-hospital mortality (n, %) | 19 (18.3%) | | 9 (16.1%) | P=0.896 |

**Legend:** Data for each patient (n=160) and each clinical event of BAL and nasal swab collection (n=189) are shown. Count data are described with numbers and percentages; distributions are described with median and interquartile range (IQR). * With reclassification based on mNGS results, total IPS cases reduced to 50 and total LRTI cases increased to 134, see PMID 38783139 and processed data files for details.
